# High clinical utility of comprehensive multi-omic molecular profiling of rare and hard-to-diagnose pediatric tumors

**DOI:** 10.64898/2026.07.25.26358936

**Authors:** Ashleigh J. Sullivan, Dong-Anh Khuong-Quang, Anita Villani, Marie Wong-Erasmus, Sarah Trinder, Loretta M.S. Lau, Paulette Barahona, Ann-Kristin Altekoester, Megan Rumford, Kimberly Dias, Chelsea Mayoh, Noemi A. Fuentes-Bolanos, Eliza K. Courtney, Sam El-Kamand, Louise Cui, Angela Lin, Scott Davidson, Kyoko E. Yuki, Nicholas Sanders, Jordan Staunton, Sophie Jessop, Shampavi Sriharan, Frank Alvaro, Antoinette Anazodo, Kanika Bhatia, Martin Campbell, Steve Foresto, Nicholas G. Gottardo, Maria Kirby, Seong Lin Khaw, Neevika Manoharan, Geoff McCowage, Andrew S. Moore, Wayne Nicholls, Matthew O’Connor, Bhavna Padhye, Anne L Ryan, Leanne Super, Paul J. Wood, Janene Davies, Colleen D’Arcy, Andrew J. Gifford, Michael Rodriguez, Katherine M. Tucker, Mark Pinese, Paul G. Ekert, Michelle Haber, Vanessa Tyrrell, Toby N. Trahair, Glenn M. Marshall, Adam Shlien, Mark J. Cowley, David S. Ziegler

## Abstract

The role of comprehensive genomic profiling for therapeutic decision-making is established in high-risk pediatric cancers, but its utility in rare and diagnostically challenging tumors is unclear. Here we report 123 non-high-risk patients enrolled in the Australian ZERO Childhood Cancer Program for diagnostic uncertainty, clinician request to address a specific molecular query, or other rare tumors. Comprehensive multi-omic profiling led to a change in diagnosis in 17.9% (22/123) of patients, with overall diagnostic utility in 35% (43/123). Molecular queries were resolved in 97.6% (40/41). Multi-omic results informed conventional management in 20.3% (25/123). Precision-guided therapy was recommended in 67.5% (83/123), and administered in 36.1% (30/83), with an objective response or prolonged (>6 months) stable disease in 88.9% of evaluable cases (16/18). Findings were confirmed in an independent cohort from the Canadian KiCS program (n=41). In conclusion, in rare and diagnostically challenging pediatric tumors, multi-omic profiling improved diagnostic accuracy and informed clinical management, supporting its integration into routine care.

## Introduction

Comprehensive molecular profiling has recently emerged as an important aspect in the management of pediatric cancer patients. Precision medicine programs for childhood cancer, including the Australian Zero Childhood Cancer Program (ZERO)^1^, the Canadian SickKids Cancer Sequencing (KiCS) Program, and others^2–5^, have been focused primarily on the identification of molecular targets to develop new precision-guided treatment strategies for high-risk patients. However, recent publications have questioned whether the potential utility in high-risk patients may extend beyond targeted therapy recommendations to include diagnostic, prognostic, and therapeutic implications^4,6–8^. Similar potential benefits have been reported when molecular profiling is performed on unselected small cohorts of pediatric patients at diagnosis^3,9^.

Very rare pediatric tumors remain poorly molecularly characterized, challenging to diagnose, and lack standardized treatment approaches. While genomic sequencing studies are often sought by clinicians to help with the diagnosis and treatment of these patients, the utility of these tests is yet to be determined. To date, there have been no studies focusing on the evaluation of the diagnostic and therapeutic utility of comprehensive molecular profiling in this patient population. To address this gap, the ZERO program^1,10^ – that integrates paired tumor-germline whole-genome sequencing (WGS), bulk transcriptome tumor analysis (WTS), and DNA methylation profiling – enrolled patients with rare, non-high-risk pediatric tumors. This cohort was independent of ZERO’s high-risk (expected cure rate <30%) cohort. Eligibility included patients with diagnostic uncertainty; for whom a specific molecular question was posed by the treating clinician; or patients with very rare tumors of uncertain prognosis and/or lacking established treatment strategies. Here, we present findings from this rare tumor cohort, alongside a validation international cohort from the KiCS program, highlighting the molecular landscape of these tumors and the impact of genomic profiling on clinical decision-making and patient outcomes.

## Results

From September 2017 to December 31, 2022, 137 consecutive non-high-risk patients were enrolled in the rare cancer cohort. Fourteen patients were excluded for analysis either due to non-neoplastic diagnoses including vascular malformations (*n=8*) or enrollment exclusively due to personal history or family history of cancer *(n=6)*. In total, 132 samples from 123 patients were included for outcome analysis (Fig. 1). These included four patients who had a second sample submitted for additional testing (i.e., second sample submitted for WGS/WTS following failed panel sequencing on initial formalin-fixed paraffin-embedded (FFPE) specimen); two patients who had two and three samples sequenced from different tumors to allow comparison to address a clinical question; and two patients who were sequenced again at relapse (Supplementary Table 1). One hundred and twelve (89.4%) patients submitted fresh, snap frozen or cryopreserved tumor samples; nine (7.3%) submitted FFPE only and two (1.6%) submitted extracted DNA from the tumor.

**Fig. 1.**
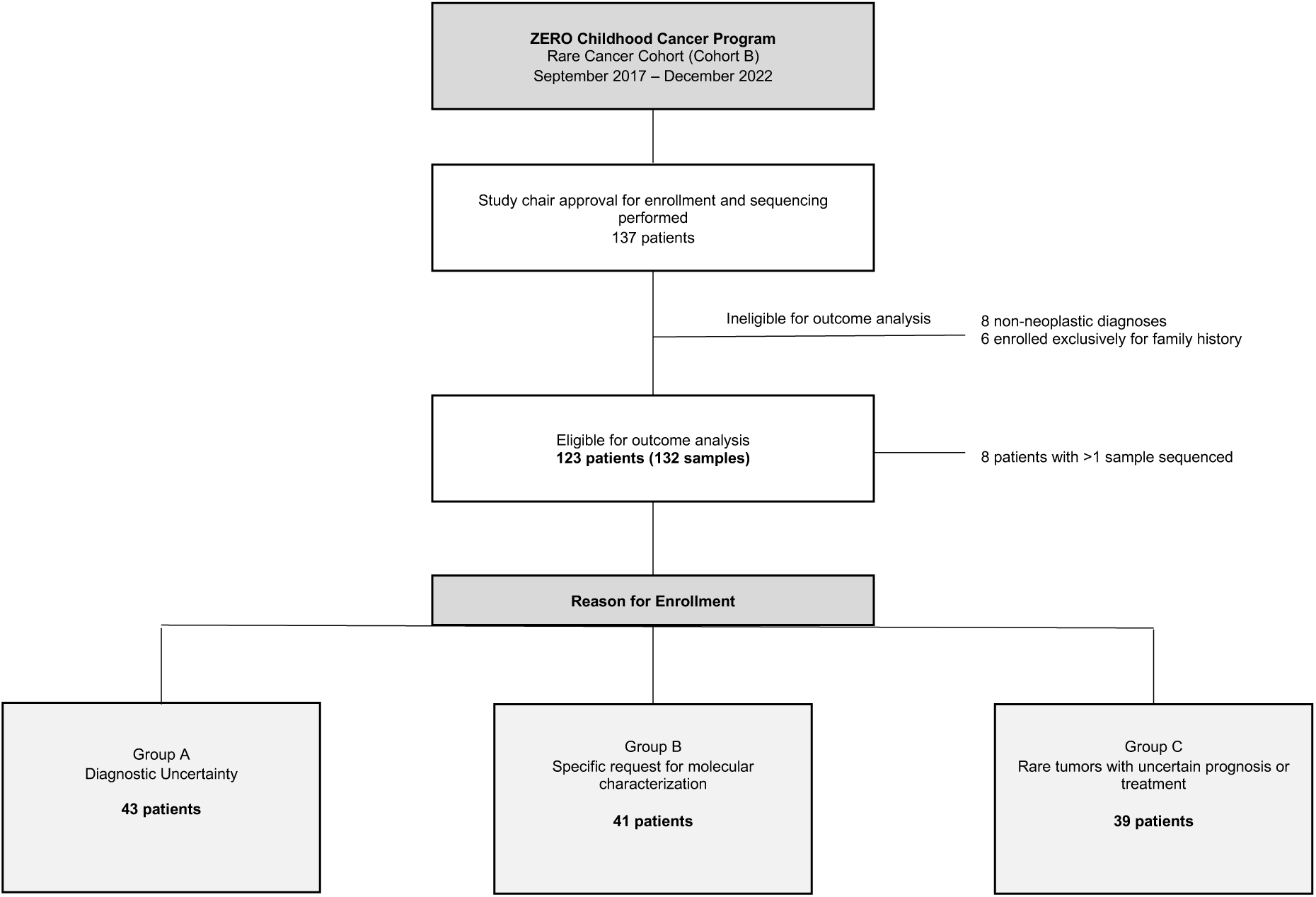
Characteristics of the cohort. A CONSORT diagram of 137 patients enrolled in the rare cancer cohort between September 2017 and December 2022. 123 patients were eligible for outcome analysis.

Patients were assigned to one of three distinct groups: Group A (diagnostic uncertainty or clarification required, *n=43*), Group B (request for testing to address a specific molecular query, *n=41*) or Group C (rare tumors with unknown prognosis, limited conventional chemotherapy options, or rare clinical scenarios, *n=39*). Patient characteristics are detailed in Extended Data Table 1. The majority (91/123; 74%) of patients were enrolled at diagnosis. As expected, given the wide range of suspected diagnoses at enrollment, genomic findings were heterogenous across the cohort (Supplementary Table 1).

Results were delivered to the treating oncologist via either multidisciplinary tumor board (MTB) discussion or a molecular report in a median time of 7.1 weeks (range 1.9–47.4 weeks). Eight patients were deceased at the time of data cutoff. Six patients were lost to follow up. Of the 115 alive patients at data cutoff, median follow up was 24.6 months (range 12.1–60.4 months).

### Comprehensive molecular profiling has high diagnostic yield in rare pediatric cancers

Multi-omic profiling led to a change in the provisional histological diagnosis in 22 out of 123 (17.9%) patients (Fig. 2a, Extended Data Tables 2 and 3). This included 20 (20/43) Group A patients, one Group B patient (1/41) and one Group C patient (1/39). Acceptance of the integrated molecular diagnosis was confirmed by the treating clinical team following ZERO recommendation in all cases. In an additional 21 (17.1%) patients, multi-omic profiling resulted in refinement in diagnosis or confirmation of a suspected, but not definitive, histopathological diagnosis (Extended Data Table 2; Supplementary Table 1). Thus overall, there was diagnostic utility in 35% (43/123) of patients (Fig. 2a).

**Fig. 2:**
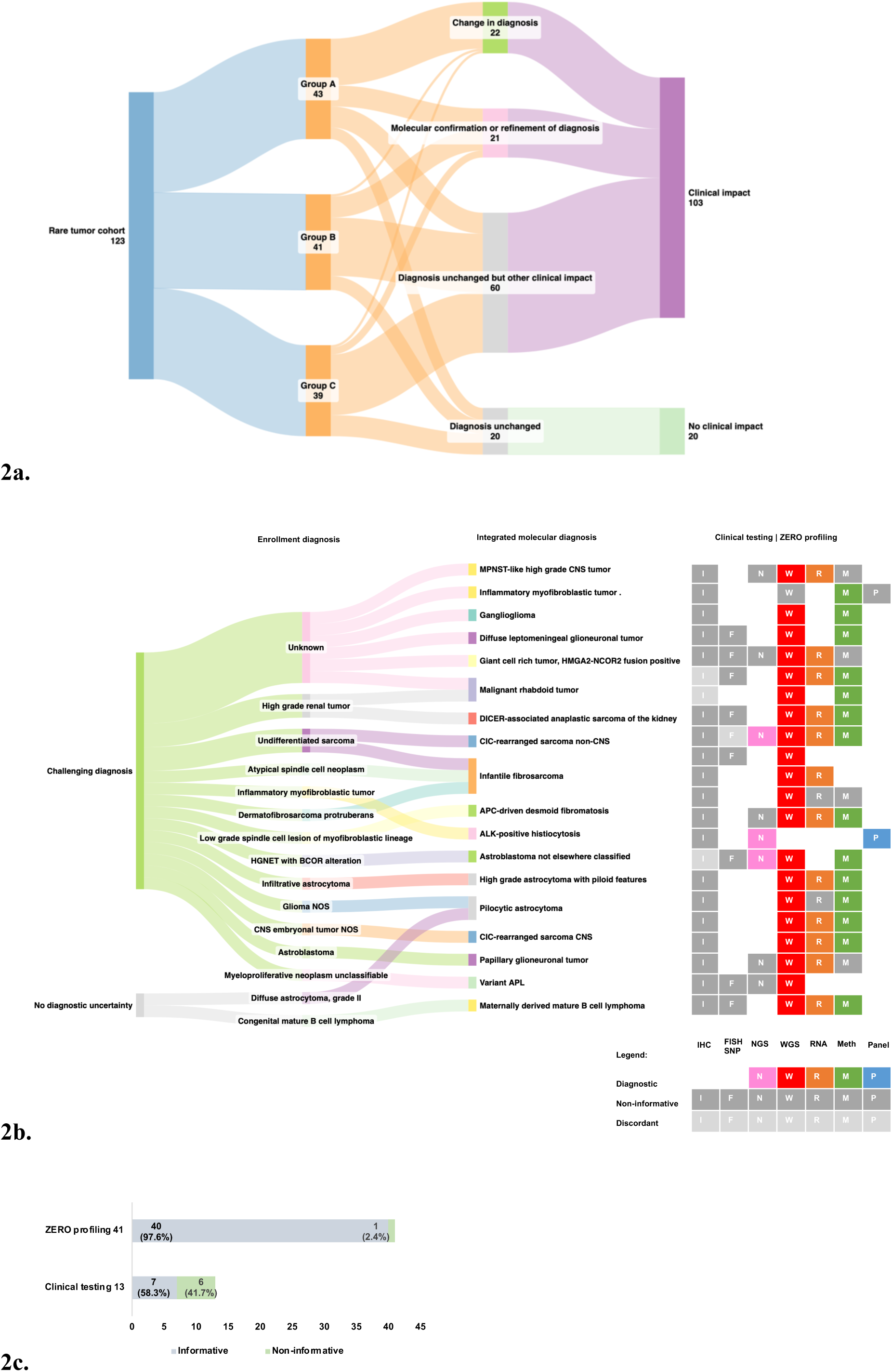
Comprehensive molecular profiling has high diagnostic yield and can address clinically relevant molecular queries. Panel A shows a Sankey diagram of 123 patients eligible for outcome analysis. Clinical impact was defined as diagnostic utility, impact on conventional oncology management, precision guided therapy (PGT) recommendation and/or treatment, prognosis, identification of an underlying pathogenic or likely pathogenic germline variant not previously known, and/or other clinician-valued benefit. Panel B shows a Sankey diagram of the 22 patients with a confirmed change in diagnosis including their enrollment diagnosis, and integrated molecular diagnosis. Clinical testing performed for each patient is listed including IHC (I), FISH or single nucleotide polymorphism (SNP) microarray (F), and targeted NGS testing (N). Non-informative results are represented by dark grey, with discordant results in light grey and informative findings in color (e.g. pink for informative NGS test). ZERO profiling performed for each patient is represented on the right, including WGS (W), WTS (R), methylation profiling (M) and panel sequencing (P). Non-informative results are represented by dark grey, with informative findings in color (red – WGS; orange - WTS; green – methylation profiling; purple – panel sequencing). Panel C shows a comparison of utility of ZERO profiling and clinical testing for 41 patients enrolled for molecular characterization. Cases in which clinical question was addressed represented by blue bars (informative). Cases where testing was not informative are represented by green bars (non-informative). Abbreviations: MPNST, malignant peripheral nerve sheath tumor; HGNET, high-grade neuroepithelial tumor; NOS, not otherwise specified; CNS, central nervous system; APL, acute promyelocytic leukemia.

Importantly, multi-omic profiling facilitated a definitive diagnosis in six patients enrolled with an unknown diagnosis and changed the diagnosis of two further patients with a firm clinical diagnosis who were not enrolled with diagnostic uncertainty (Extended Data Table 3). In this subgroup of 22 patients whose diagnosis changed (20 Group A, one Group B, one Group C), WGS ± WTS contributed to the revised diagnosis in 20 out of 22 cases (Fig 2b; Extended Data Table 3). The remaining two cases had their diagnosis clarified based on a targeted sequencing panel (n=1), where only an FFPE sample was available, or by methylation profiling (n=1).

We next considered which components of the multi-omic platform contributed to the diagnostic utility (Fig 2b). Importantly, a combination of different molecular tests provided diagnostic utility in 17/21 cases. Fourteen of the revised diagnosis cases had both WGS and WTS (WGTS) performed. In five of these cases (35.7%), WTS was essential to identify or confirm driver alterations (e.g *EML4::NTRK3* fusion identified only by WTS in an infantile fibrosarcoma). In a further seven cases (50%), WTS provided additional supportive evidence or orthogonal validation (e.g. confirming loss of NF1 function through the absence of the mutated allele in a high-grade astrocytoma). Concordance of molecular and methylation results was seen in 14 of the 18 (77.8%) patients with methylation profiling performed. Tumor classification by methylation arrays was essential to establish a diagnosis in one patient with an uncertain histological diagnosis and low tumor purity (24%), where WGS yielded no reportable findings: a high calibrated score (0.99, MNP classifier v12.8) supported inflammatory myofibroblastic tumor, which was accepted as the final diagnosis by the clinical team. Thus, all components of the comprehensive molecular profiling platform contributed to a change or refinement in the initial provisional diagnosis across the entire Rare Tumor Cohort (Fig 2b).

We next evaluated which genomic aberrations provided diagnostic utility (Extended Data Table 3). Diagnostic genetic alterations varied across different tumor types, however, structural variants were the most common alteration providing diagnostic utility, contributing to a change in provisional histological diagnosis in 13 of the 22 patients. Germline WGS contributed to the diagnosis in two patients and led to the identification of previously unrecognized cancer predisposition conditions within the broader family. In one case (zccs1519), a child enrolled with a low-grade spindle cell tumor with nuclear beta-catenin positivity but negative *CTNNB1* sequencing was enrolled for diagnostic clarification. Paired tumor-germline sequencing revealed a heterozygous germline pathogenic frameshift *APC* variant, somatic loss of 5q22.2 (including *APC*), low *APC* RNA expression, and a methylation profile consistent with desmoid fibromatosis, which together confirmed the correct diagnosis. In another case (zccs1342), a high-grade renal tumor with retained BRG1 on immunohistochemistry was shown to harbor a germline pathogenic frameshift *SMARCA4* variant and a second somatic hit, resulting in complete *SMARCA4* loss. This, along with a supportive methylation classification, confirmed a diagnosis of malignant rhabdoid tumor and highlighted the limitations of using immunohistochemistry (IHC) screening alone. Similarly, three additional patients with a revised integrated diagnosis had false positive or false negative IHC or fluorescence *in situ* hybridization (FISH) results, and their diagnoses relied on multi-omic analysis (Fig. 2b, Extended Data Table 3).

#### Diagnostic yield in Group A (diagnostic uncertainty cohort)

Among the 43 Group A patients, comprehensive multi-omic profiling combined with histopathology resolved the diagnostic uncertainty in 46.5% (20/43; Extended Data Table 3). That is, revised diagnoses were primarily seen in patients enrolled due to diagnostic uncertainty, with 20 of the 22 (90.9%) revised diagnosis cases occurring in Group A (diagnostic uncertainty cohort; n=43), thus highlighting the value of genomic testing in this setting (Fig. 2a). In contrast, of the nine patients in this group who underwent concurrent clinical molecular testing on various clinically accredited small next-generation sequencing (NGS) panels, only three had informative results, while the remaining six had non-informative or discordant findings (Extended Data Table 3).

An additional nine Group A patients had their diagnoses refined or confirmed (Extended Data Table 2) through multi-omic testing, bringing the overall diagnostic utility to 67.4% (29/43) in this group. Routine testing methods were at the discretion of the enrolling center, and included provisional histopathology with IHC and/or FISH, cytogenetics and/or clinically accredited targeted NGS. An expert histopathology review had been sought in at least 15 of 43 (34.9%) Group A cases. Multi-omic profiling in conjunction with discussions with the local clinical team led to a revised diagnosis in 7 of these 15 patients.

### Multi-omic profiling provides definitive answers to real-world clinical molecular queries

Multi-omic profiling effectively addressed diverse clinician queries in Group B (request for testing to address a specific molecular query), resolving 97.6% of cases (40/41). These included requests for characterization or confirmation of a specific fusion or mutation (*n=27*), or pathway activation (*n=3*), molecular confirmation of a favored diagnosis (*n=1*), molecular subtyping (*n=6*) or other molecular queries (*n=4*), such as the relationship between primary and secondary tumors or evidence of mismatch repair deficiency. In all 39 cases with fresh frozen or cryopreserved tissue undergoing WGS and where possible, WTS, the platform addressed the clinician queries (Fig. 2c). Of two cases where only FFPE samples were available and underwent panel sequencing, one was non-informative (Fig. 2c). Concurrent clinical laboratory testing to address the enrolment query was performed in 13 of 41 Group B cases; in six, the clinical test targeting the specific molecular alteration was non-informative (Fig. 2c). In four additional cases, clinical tests, including FISH and IHC, produced false positive or false negative results compared to molecular (Supplementary Table 1; zccs1223, zccs1410, zccs1469, zccs1473). In addition to answering the specific clinical question, the multi-omic platform provided molecular confirmation or refinement of the diagnosis in 24.4% of Group B patients (10/41). Thus, multi-omic testing was highly efficient at resolving specific clinical queries, and overall provided more utility than available clinical tests.

### Comprehensive multi-omic profiling improves clinical management of patients with rare pediatric tumors

We next assessed the clinical impact of multi-omic profiling on patient outcomes. Notably the Rare Tumor cohort overall had a favorable prognosis (Extended Data Fig. 2), with a 2-year progression free survival (PFS) and overall survival (OS) of 82.9% and 93.2% respectively, in contrast to patients enrolled in the high-risk cohort^1^. We therefore sought to understand the clinical impacts of multi-omic profiling in this unique group of patients beyond providing diagnostic information.

Molecular results directly informed at least one component of conventional oncology treatment in 25 of 123 patients (20.3%), leading to adjustments in chemotherapy (*n=16*), radiotherapy (*n=6*), hematopoietic stem-cell transplant (HSCT) (*n=2*), and surgical decisions (*n=7*) (Fig. 3). These changes included de-escalation of prior planned chemotherapy (*n=4*), radiation omission or dose reduction (*n=3*), and avoidance of high-risk or mutilating surgery (*n=4*).

**Fig. 3:**
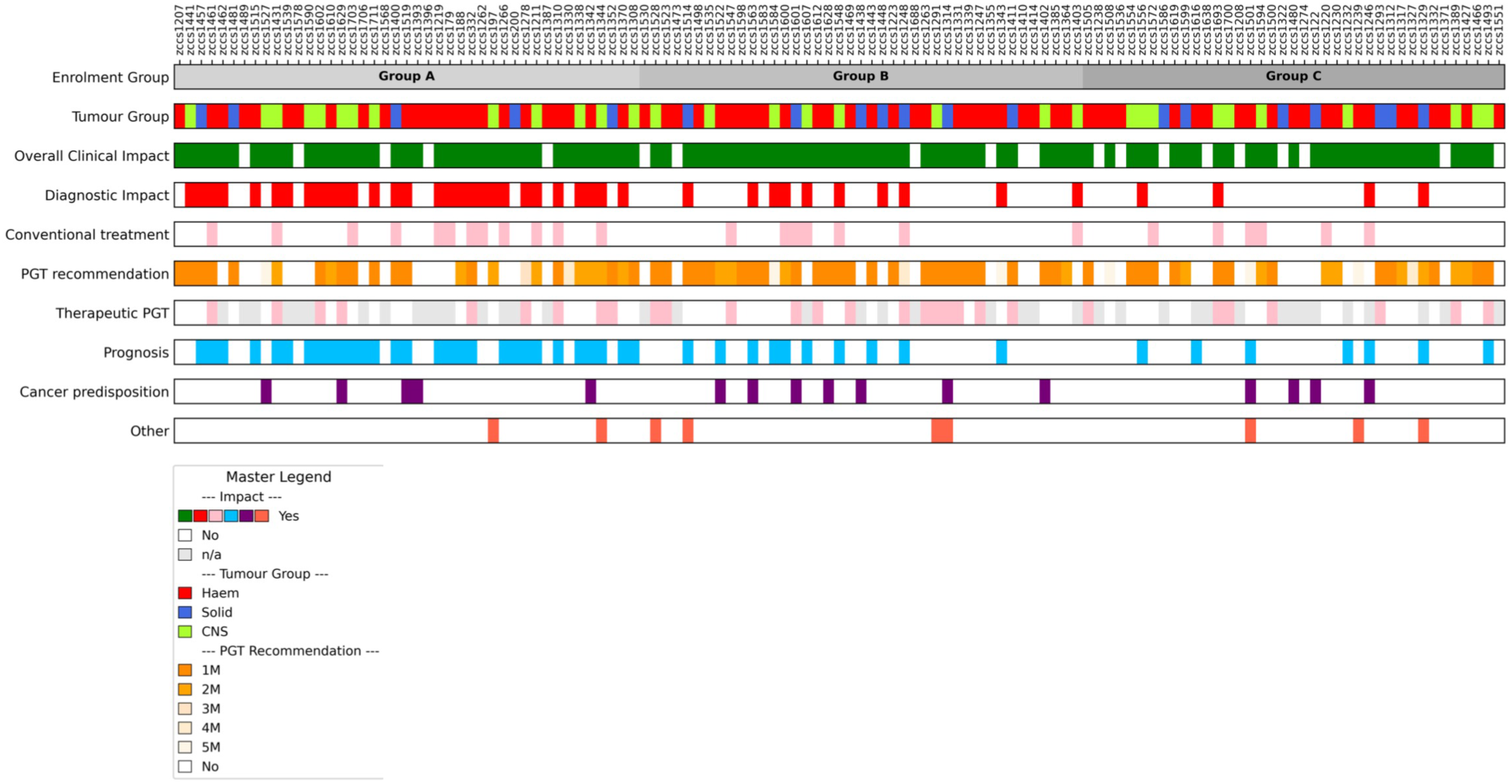
Comprehensive molecular profiling improves clinical management of patients with rare cancers. An oncoprint demonstrating clinical impact for all 123 ZERO patients grouped by their reason for enrollment (Group A: diagnostic uncertainty or clarification required; Group B: request for testing to address a specific molecular query; Group C: other rare tumors including tumors with unknown prognosis, limited conventional chemotherapy options, or rare clinical scenarios). Patients with an overall clinical impact are represented by green with the individual impacts represented below: diagnostic impact (either molecular confirmation, molecular refinement of diagnosis, or change in the provisional histopathological diagnosis; red), change in therapeutic conventional oncology management (informed decision to give, adjust or omit chemotherapy, radiation, surgery and/or HSCT; pink), PGT recommendation (orange; shades indicate tier of recommendation) and PGT therapy (pink), prognosis (blue), identification of a previously unknown germline finding leading to recommendations in surveillance and/or cascade testing (purple), or other clinical impact (dark pink). Overall, 82.9% of participants had at least one clinical impact (green). Abbreviations: PGT, precision guided therapy; haem, hematological; CNS, central nervous system.

Multi-omic profiling identified precision-guided therapy (PGT) recommendations in 67.5% (83/123) of patients, of which notably the vast majority 87.9% (73/83) were Tier 1M (clinical evidence in the same tumor type) or Tier 2M (clinical evidence in another tumor type).^1,10^ Of these 83 patients, 36.1% received the recommended PGT (n=30). Twelve of the 30 patients were not evaluable for PGT response due to insufficient data availability (n=5), inadequate clinical or radiological data to perform formal response assessment (n=3), conventional chemotherapy co-administered (n=3) or no measurable disease at baseline (n=1). Among the 18 patients evaluable for objective response to therapy, 88.9% (16/18) demonstrated objective responses or prolonged stable disease of >6 months (Fig. 4a, 4b), with a median treatment duration of 10.2 months (range 1.2–26.6 months). Complete or partial responses were observed in 55.6% (10/18), with a median duration of response of 14.5 months. The overall disease control rate was 88.9% (16/18).

**Fig 4:**
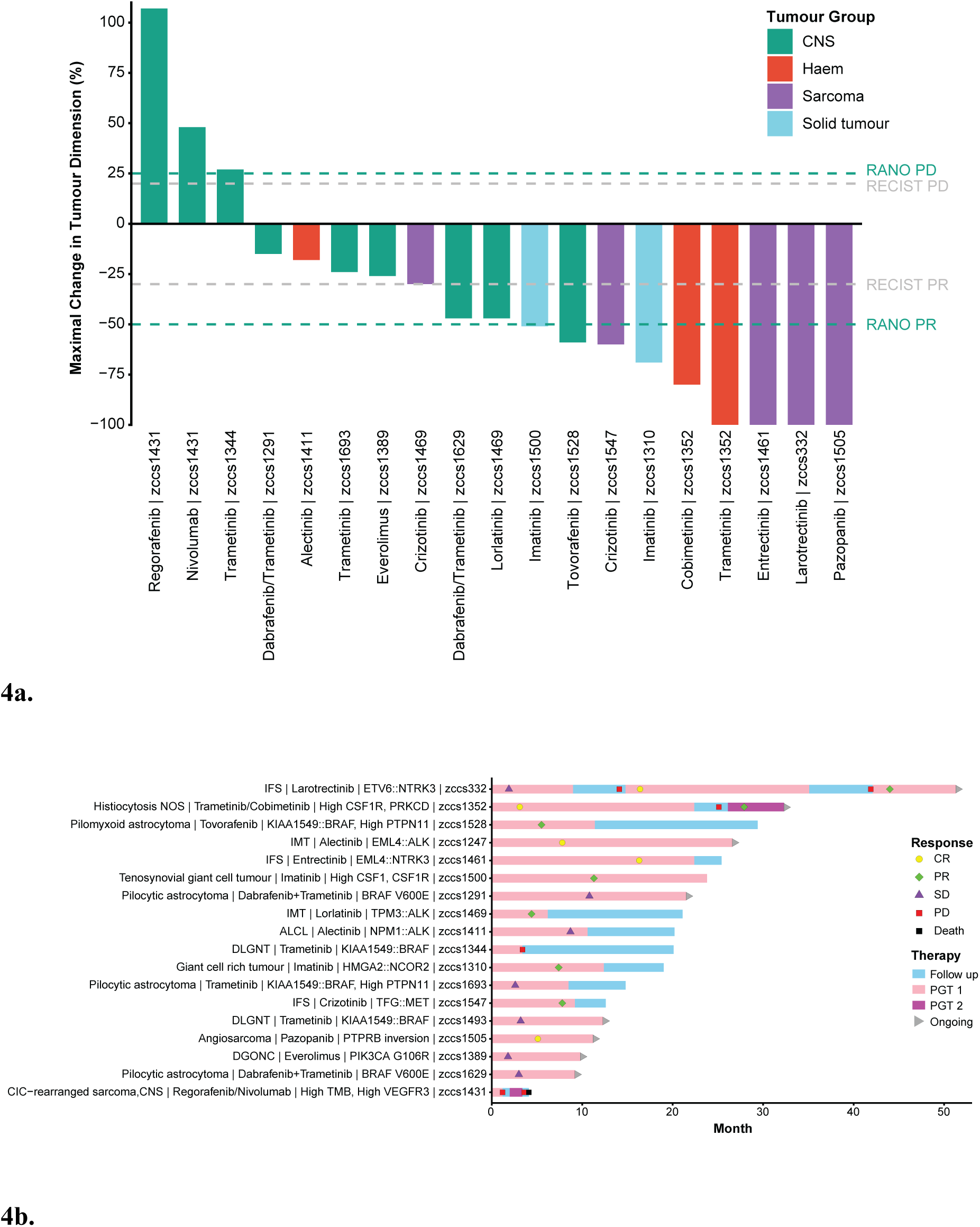
PGT in rare tumors leads to durable objective clinical benefit. Panel A shows a waterfall plot demonstrating maximal change in tumor size (%) of all patients with measurable disease receiving PGT who met eligibility for response assessment. Treatment response was evaluated using RANO criteria for CNS tumors, and RECIST or PERCIST criteria for solid tumors. The grey dotted lines at 20% and -30% delineate the category of response according to RECIST criteria (≥ 20%, PD; 20 to -30%, SD; -30% or lower to -99%, PR; -100% CR). The green dotted lines at 25% and -50% delineate the category of response according to RANO criteria (≥ 25%, PD; 25 to -50%, SD; -50% or lower to -99%, PR; -100% CR). Colored bars indicate diagnostic category. One patient with a multifocal bone histiocytic disorder (zccs1352) appears twice in the waterfall plot, as they received two different PGT: trametinib and cobimetinib; trametinib was ceased due to adverse effects. Panel B shows a swimmer plot demonstrating all 18 patients who received PGT and met eligibility for response assessment including diagnosis, targetable alteration and therapy administered. Denoted on the left is final diagnosis; drug(s) received with (+) to indicate PGT given in combination and (/) to indicate PGT given separately; targetable mutation; study ID. Light pink bars indicate period of patient receiving a PGT agent, purple bars indicate period of patient receiving different second PGT agent. Blue bars indicate period of follow up for patient while not receiving PGT. One patient (zccs1500) did enter follow up ten days prior to data cut-off however the off treatment follow up time period is too brief to be reflected as a blue bar. Abbreviations: ALCL, anaplastic large cell lymphoma; CNS, central nervous system; DGONC, diffuse glioneuronal tumor; DLGNT, diffuse leptomeningeal glioneuronal tumor; IFS, infantile fibrosarcoma; IMT, inflammatory myofibroblastic tumor; NOS, not otherwise specified; TMB, tumor mutational burden; PGT, precision guided therapy.

Molecular findings also informed prognosis in 37.4% (46/123) of cases through either informing diagnosis or revealing a molecular finding with prognostic impact (Fig. 3). Further, germline pathogenic or likely pathogenic variants in cancer predisposition genes were observed in 20.3% (25/123), including 17 patients where the finding was not known prior to enrollment (13.8%; 17/83). This prompted cancer genetics consultation, individualized cancer surveillance and/or cascade testing. In 11 of these 17 newly identified cases, variants had a well-established association with the tumor (tumor-cognate), while six involved secondary unexpected findings requiring cancer genetics consultation and/or cancer surveillance and cascade testing.

Additional clinical utility was observed in nine patients, including facilitating trial enrollment, identifying precise fusion breakpoints to facilitate minimal residual disease marker development for leukemia disease monitoring, personalizing surveillance and, in one exceptional case, detecting maternofetal transmission of mature B-cell lymphoma in a neonate.

Importantly, patients in Group C (other rare tumors including tumors with unknown prognosis, limited conventional chemotherapy options, or rare clinical scenarios) were also observed to benefit from genomic sequencing (Fig. 3). In 30.8% (12/39), results either informed chemotherapy treatment or led to initiation of PGT, and prognostic insights were revealed in 17.9% of patients (7/39). In addition, sequencing led to diagnostic impact in four (10.3%; 4/29) Group C patients including one patient with a change in diagnosis. Thus, molecular findings collectively influenced conventional oncology management, guided therapy selection, informed prognosis, identified previously unrecognized germline variants, and provided other clinician-valued benefits across all enrollment groups. Overall, 82.9% (102/123) of all patients experienced at least one positive clinical impact.

### Comprehensive molecular testing identifies novel genomic alterations in rare and hard to diagnose pediatric tumors

Multi-omic profiling revealed novel or extremely rare genomic findings in 12/123 patients (9.8%) (Table 1). Genomic insights included novel findings within specific cancer types, such as previously unreported variants in children, and new fusion partners^11–13^. For example, a novel *ALK* fusion was identified, with a previously unreported *FXR1* fusion partner, in an infant high-grade glioma (zccs1535). In a patient with choroid plexus carcinoma (zccs1584), sequencing revealed a pathogenic truncating somatic *SUFU* mutation, representing a previously unreported potential therapeutic target in this disease.

**Table 1:** Summary of novel and rare molecular findings. Novel indicates previously unreported variants or fusion partners in a specific cancer type. Ultra-rare indicates findings that have been rarely reported in the literature previously^11,12,14,15,28–31^.

| ID | Diagnosis and Molecular finding | Subcategory | Comments and/or publication |
| --- | --- | --- | --- |
| zccs1584 | Choroid plexus carcinoma<br>SUFU: c.1309G>T (p.Glu437Ter) | Novel | Not previously reported clinically. |
| zccs1291 | Pilocytic astrocytoma<br>BRAFV600_K601delinsE | Novel | Extremely rare BRAF exon 15 variant not reported in pediatric cancer. |
| zccs1535 | Infant-type hemispheric glioma<br>FXR1::ALK fusion | Novel | Novel ALK fusion partner. |
| zccs1330 | Myofibroma<br>MARCKS::FRK fusion | Novel | Not previously published in this tumor type. |
| zccs1703 | High grade astrocytoma with piloid features<br>Germline PTPN11 mutation | Novel | Novel presentation of this tumor in a patient with Noonan Syndrome - case published<sup>12</sup>. |
| zccs1498 | Plexiform neurofibroma<br>Somatic NF1 loss | Novel | Somatic NF1 loss in sporadic plexiform neurofibroma not previously reported. |
| zccs1248 | Langerhans cell histiocytosis<br>BRAF: c.1511_1517+2dup | Ultra-rare | Rare somatic splicing variant reported only twice previously in single site LCH. |
| zccs1310 | Giant cell rich tumor, HMGA2-NCOR2 positive<br>HMGA2::NCOR2 fusion | Ultra-rare | Patient included in pediatric case series<sup>11</sup>. |
| zccs1514 | Malignant cutaneous myeloid/monocytic neoplasm<br>TPM3::ALK fusion | Ultra-rare | Rare in congenital myeloid neoplasm; previously reported in other tumor types. |
| zccs1610 | Infantile fibrosarcoma<br>BRAFV600E mutation | Ultra-rare | Case-series of 14 patients with BRAF alterations and histology overlapping with IFS<sup>28</sup>. |
| zccs1400 | Variant acute promyelocytic leukemia<br>TBLXR1-RARB fusion | Ultra-rare | Rare pathognomic fusion for variant APL with therapeutic significance, reported 5x previously<sup>14-15</sup>. |
| zccs1590 | Astroblastoma, not elsewhere classified<br>EWSR1-BEND2 fusion | Ultra-rare | Reported rarely in this tumor type<sup>29-31</sup>. |

Beyond novel findings, additional cases also expand the literature on rare pathognomonic mutations and fusion. For example, a 6-year-old boy with hyperleukocytosis and dysplastic promyelocytes was initially diagnosed with a myeloproliferative neoplasm, unclassifiable (zccs1400) after inconclusive standard testing. WGTS identified the rare but pathognomonic *TBL1XR1::RARB* fusion, confirming a diagnosis of variant acute promyelocytic leukemia (APL), with important prognostic and therapeutic implications (i.e., AML-directed chemotherapy instead of upfront HSCT)^14,15^. In another case, a difficult to diagnose parietal tumor with a provisional histological diagnosis of astroblastoma-like tumor was found to harbor the rare *SLC44A1::PRKCA* gene fusion: highly specific for papillary glioneuronal tumors, informing diagnosis and a favorable 5-year PFS exceeding 80%^16,17^.

The multi-omic approach revealed critical findings that may have been missed with either WGS or WTS alone. Indeed, for 28 of the 81 patients who underwent combined WGTS (34.5%), the WTS was essential to identify relevant molecular findings influencing diagnosis, therapeutic opportunity, and management (Supplementary Table 1). For example, WTS was critical when complex structural variants or chromothripsis obscured identification of a driver fusion by WGS (i.e., chromothripsis of chromosome 5 resulting in an in-frame *TPM4::ALK* fusion), or in a low tumor purity sample where the alteration was missed on 90x WGS due to insufficient read depth (i.e., *GATA1*:c.150_151insTG in AML detected at 2% allelic fraction by WTS). In a further 38 cases (46.9%), WTS provided orthogonal validation of the WGS findings and/or provided additional supportive evidence for the pathogenic role of the genomic alteration identified, such as loss of expression of mutated tumor suppressor genes such as *NF1*, *TP53,* and *SMARCB1*, consistent with nonsense mediated decay.

### Clinical utility replicated using a Canadian pediatric cancer cohort

The utility of the comprehensive ZERO platform may be influenced by the local availability (or lack-there-of) of conventional clinical tests. We therefore sought to validate our findings with an independent international cohort using our multi-omics analysis pipelines. Forty-one patients enrolled in the Canadian KiCS precision medicine program met inclusion criteria for groups A (n= 22), B (n=11), and C (n=8). This cohort showed similarly high rates of diagnostic yield and impact on clinical management, compared to the ZERO cohort (overall 73.1%, Extended Data Table 4). A revised diagnosis was made in 12 of 22 (54.5%) patients in Group A, and molecular refinement was provided for two further patients, with additional diagnostic utility emerging from Group B and C (Extended data Fig 3). WGS ± WTS answered specific molecular questions for all 11 (100%) patients in group B. These findings were not reported in the 5 patients who underwent concurrent clinical testing (n=5). Collectively, across all groups, in the KiCS cohort, comprehensive profiling impacted conventional oncology treatment in 24.3% and prognosis in 43.9% of patients, respectively (compared with 20.3% and 37.4% in the ZERO cohort).

All genomic findings that informed diagnosis or other aspects of clinical utility were concordant between the KiCS and ZERO programs, with the exception of two cases with *EGFR* internal tandem duplication (ITD), not confidently called on original panel-based testing by KiCS. One of these cases, an infant with a flank/extremity soft tissue tumor originally called an undifferentiated sarcoma, would now be characterized as an infantile fibrosarcoma-like neoplasm. The second case, originally diagnosed in 2021 as a mesenchymal hamartoma of the aryetnoid in an infant, would now be characterized as a spindle cell tumor with EGFR-ITD, low-grade histology and lipofibromatosis-like pattern on morphology. These cases reflect evolution of both genomic technologies and analysis pipelines for variant detection and more recent reclassification^18,19^ of rare pediatric tumor entities. Overall, ZERO’s findings of clinical utility were reproducible in an independent, international clinical cohort of rare and histologically uncertain pediatric cancers.

## Discussion

Previous large childhood cancer precision medicine studies have established the feasibility and potential benefits of comprehensive genomic profiling platforms in high-risk pediatric cancer cohorts^1,5,7,8,20^. However, the role of precision medicine platforms for pediatric patients with rare and/or hard to diagnose tumors has not been reported. This study demonstrates, for the first time, that the use of comprehensive genomic sequencing has major clinical impact by improving diagnostic accuracy, personalizing conventional management decisions (including the de-escalation of therapy), and identifying effective PGT options.

Genomic findings influenced diagnosis in 35% of patients in the entire cohort either by a formal change in the provisional histopathological diagnosis, molecular confirmation of a suspected diagnosis, or diagnostic refinement. Of the patients enrolled with diagnostic uncertainty (Group A), the comprehensive genomic testing provided diagnostic insight in 67.4% of patients, highlighting a subgroup of patients in whom clinical benefit is likely to be high. Importantly, even in the 14 diagnostic uncertainty (Group A) patients in whom genomic testing did not provide diagnostic insights, clinical impact was still observed in 64.3% (9/14). In this cohort, diagnosis changes were diverse, with WTS and/or methylation profiling essential in 6 of the 22 revised diagnosis cases (Fig. 2b). Nearly a quarter of cases required WTS to identify or confirm the clinical impact of the key molecular finding, and an additional 30.9% benefited from orthogonal validation or supportive WTS data. This underscores the value of a multi-omic approach and supports routine integration of WTS in precision medicine platforms for rare tumors^3,21^.

Patients in Group B, enrolled for specific molecular queries, also benefited from the comprehensive approach, which resolved all cases where WGS ± WTS was performed. Notably, only 31.7% had received clinical testing to address the enrolment question, likely reflecting real-world barriers such as test availability and reimbursement, access issues, and clinician genomic literacy. Even in cases where concurrent clinical testing was performed, the results proved less informative than those obtained through the comprehensive ZERO platform. This highlights the value of a comprehensive platform compared with clinical testing, especially in pediatric oncology where limited biopsy material must be efficiently utilized across multiple tests and commercially available testing is often limited to screening panel-based tests targeting common alterations.

Independent of diagnostic utility, 20.3% of the cohort experienced changes to conventional management based on multi-omic results. This impact was seen not only in patients with a change in diagnosis but across the entire cohort (Fig. 3). Given that most patients (74%) were enrolled at diagnosis and generally had a favorable prognosis, the implication of reducing potentially toxic therapy suggests meaningful impact, with 10 patients receiving less treatment based on molecular results.

Most of the cohort received a PGT recommendation (67.5%). Whilst this is consistent with our own^1,10^ and other studies^2–5^ that have reported on the therapeutic opportunity of molecular profiling in high-risk pediatric cancers, this is of specific relevance for rare tumors as in many cases there may be no established conventional treatment and PGT may be an appropriate first-line therapy in the upfront setting. This may explain the relatively high PGT uptake^1,4^ of 36.1%, again highlighting a unique potential for clinical impact in this patient group. It is also notable that the vast majority of recommendations in this cohort were high Tier 1 or 2M, highlighting the potential clinical impact, and correlating with high clinical benefit.

This study revealed greater clinical impact than in some other precision medicine pediatric oncology reports. While previous studies have reported diagnostic impact rates ranging from 3–16.6%, our findings demonstrate a higher level of diagnostic utility in this rare tumor cohort^4,7,8,22^. For example, the Genomic Medicine Sweden Childhood Cancer project, enrolling all children diagnosed with a primary or relapsed solid malignancy in Sweden, reported diagnostic refinement in 56% of upfront cases, but only one patient had a change in diagnosis^3^. Similarly, Trotman et al. observed a 16.6% diagnostic impact in newly diagnosed solid and central nervous system (CNS) tumors, with just four patients with a change in provisional diagnosis^2^. In contrast, our cohort had a higher rate of diagnostic change and refinement, likely due to targeted enrollment of patients with diagnostic uncertainty. Recent publications have also noted that broadly in pediatric cancer, potential impacts of genomic sequencing may extend beyond PGT^2,4,9^. In a UK cohort of newly diagnosed patients, 7% of patients had a change in conventional oncology management^9^. These results contribute to the growing evidence supporting routine molecular sequencing in pediatric oncology.

These results should be interpreted in light of the study’s limitations. The ZERO program reflects the Australian healthcare context and is influenced by variable provision of standard of care (SOC) testing, including non-universal access to clinical targeted NGS panel testing across sites and funding constraints. However, validation of these findings in an independent international cohort of similar patients supports their broader relevance. Additionally, genomic literacy among clinicians has evolved since the inception of this program, and the interpretation of the molecular findings was based on published literature available at the timepoint of each patient’s MTB discussion. Furthermore, in considering the applicability to clinical practice, additional research exploring the cost-effectiveness of this approach as well as patient reported outcomes is essential.

In conclusion, this study demonstrates the high clinical utility of comprehensive multi-omic profiling in hard-to-diagnose and rare pediatric tumors. This provides a strong rationale and foundation for implementation into clinical practice to facilitate provision of personalized genomic-informed care for children with rare and hard-to-diagnose cancers.

## Methods

### Trial oversight

The PRISM (PRecISion Medicine for Children with Cancer) study (NCT03336931) was a multi-center prospective observational cohort study conducted in the Australian ZERO Childhood Cancer Precision Medicine Program and was opened from September 2017 in all eight pediatric oncology centers in Australia. The study was conducted in accordance with Good Clinical Practice guidelines and the Declaration of Helsinki and approved by the Hunter New England Human Research Ethics Committee of the Hunter New England Local Health District in Australia (reference no. 2019/ETH00701).

The KiCS study was opened in April 2016 for patients at The Hospital for Sick Children in Toronto, Canada, but also enrolled patients nationally and internationally. The study was approved by The Hospital for Sick Children Research Ethics Board.

### Patients and samples

Patients aged <21 years with a rare pediatric tumor with eligibility as defined above, were eligible for enrollment in the PRISM rare cancer cohort after approval by the study chair. A patient was deemed eligible when the following criteria were satisfied: study chair approval for enrollment, both tumor and germline samples received at Children’s Cancer Institute (Sydney, Australia), and sufficient DNA could be extracted for sequencing. Tumor tissue was received fresh, snap-frozen or cryopreserved. An FFPE tumor sample was accepted with approval from the study chair if this was the only sample available, and panel sequencing was deemed likely to be effective. Clinical and demographic data at registration and outcome data at subsequent follow-up time points were collected.

For both Australian and Canadian cohorts, after review by at least two investigators, enrolled patients were retrospectively assigned into three groups based on the reason for enrollment by their primary clinician: diagnostic uncertainty or clarification (Group A), request for further molecular characterization with a specific query (Group B), or other rare tumors with limited conventional chemotherapy options, unknown prognosis or rare clinical scenario (Group C).

### Trial procedures

WGS (paired tumor-germline to >90x and >30x depth, respectively) was performed on all patient samples, except where there was insufficient tumor DNA for WGS or where only an FFPE tumor sample was available. Illumina TruSight^TM^ Oncology 500 targeted panel sequencing was performed on these patient samples. All non-FFPE tumor samples underwent WTS to a depth of 100M reads when adequate RNA was available and not degraded. DNA methylation profiling was performed in CNS tumors, suspected sarcomas and selected other tumors using Illumina Infinium EPIC array (V1) and data were analyzed using the German Cancer Research Centre (DKFZ) Molecular Neuropathology (MNP) classifier for CNS tumors (v11b4, 12.3, 12.5)^23^ or Sarcomas (v12.2)^24^. The analytic pipelines for molecular profiling and variant curation for WGS, WTS, methylation profiling and targeted panel sequencing and the ZERO reporting process including tiers for therapy recommendation have been described previously^1,10^. The KiCS program procedures have been described previously^4^. In brief, real-time results for KiCS patients were reported from an 864-gene paired tumor-germline cancer panel and WTS; matched WGS data were not actively analyzed at the time of reporting. Cases eligible for Groups A–C were identified, and WGS ± WTS data were shared and independently analyzed via the ZERO analysis pipelines.

### Outcomes

Patients with a non-neoplastic diagnosis, including vascular malformations, or patients enrolled solely for germline analysis due to personal or family history were excluded from outcome analysis.

Primary outcomes included the proportion of patients receiving a recommendation for 1) a change in diagnosis or 2) a change in treatment. Secondary outcomes included 1) the proportion of tumor samples found to have molecular alterations resulting in a change in diagnosis and/or therapy, including chemotherapy, radiation, HSCT, 2) the turnaround time for report, 3) the proportion of patients who receive personalized therapy and 4) the response rate to recommended therapy. Further outcomes were explored using secondary analysis including 1) the proportion of cases where there was molecular refinement or confirmation of a favored diagnosis, 2) the ability of the universal ZERO multi-omic platform to address specific molecular questions, 3) patient OS and 4) patient PFS.

All recommendations for change in diagnosis, treatments (including conventional and experimental treatments), as well as uptake of recommendations and treatment responses were recorded prospectively. The treating oncologist’s opinion of the impact on clinical management was also recorded. Extended clinical impact data were obtained by chart review, from the sample management system, Labmatrix® (IQVIA Laboratories), and/or from the treating clinician if required. All treatments and response to treatment were collected prospectively in Labmatrix and verified through medical record review. Disease responses were evaluated using the Response Evaluation Criteria in Solid Tumors (RECIST, v1.1)^25^, Positron Emission Tomography Response Criteria in Solid Tumors (PERCIST)^26^ or Response Assessment in Neuro-Oncology criteria^27^. Follow-up data cut-off was set at 31 December 2023.

Primary and secondary outcomes of patients in the KiCS validation cohort were reported based on the analysis and interpretation at the time of initial case review by the KiCS program. Additionally, there was an independent analysis and interpretation by the ZERO team using the analytic pipeline described above.

### Statistical analysis

Descriptive statistics were used to describe baseline demographics of the patient cohort, and no statistical method was used to predetermine sample size. The Kaplan-Meier method was used to analyze survival (OS and PFS) where censoring was random. No statistical comparisons between cohorts were conducted. Statistical analyses were performed using the GraphPad PRISM 10 software. Kaplan-Meier survival curves were generated using custom code, developed with computational assistance; all analyses were performed and verified by the authors. Sankey plots were created using SankeyMATIC.

## Data Availability

All data produced are available online at the European Genome-Phenome Archive under accession numbers EGAS00001008471, EGAS00001004572, EGAS00001007937.

## Data availability

WGS, RNAseq and methylation data generated by this study are available from the European Genome-phenome Archive under accession numbers EGAS00001008471, EGAS00001004572, EGAS00001007937. A Supplementary Data file contains individual patient demographic data, integrated diagnoses, sequencing performed including RNASeq impact and reportable molecular aberrations.

## Acknowledgements

The Zero Childhood Cancer National Precision Medicine Program has been funded through a joint initiative of the Australian Government’s Medical Research Future Fund’s Emerging Priorities and Consumer Driven Research scheme and the Minderoo Foundation (2020-2025). The New South Wales State Government, Australian Federal Government and the Australian Cancer Research Foundation provided funding for the establishment of infrastructure to support the Zero Childhood Cancer National Precision medicine program. Funding from the Kids Cancer Alliance, Cancer Therapeutics Cooperative Research Centre, supported the initial development of the precision medicine program (2015); Tour de Cure supported tumor biobank personnel; and the Lions Kids Cancer Genome Project, a joint initiative of Lions International Foundation, the Australian Lions Children’s Cancer Research Foundation (ALCCRF), the Garvan Institute of Medical Research, the Children’s Cancer Institute and the Kids Cancer Centre, Sydney Children’s Hospital provided funding to perform WGS (2016– 20) and for key personnel, with thanks to J. Collins for project governance and advocacy. The Cure Brain Cancer Foundation supported the RNA sequencing of patients with brain tumors (2016–8); the Kids Cancer Project supported molecular profiling and molecular and clinical trial personnel (2014–20); and the University of New South Wales and W. Peters provide personnel funding support. The 2018 Priority-Driven Collaborative Cancer Research Scheme (APP1165556), co-funded by Cancer Australia and My Room, Hartwig Medical Foundation and Australian BioCommons supported methods development, personnel funding and molecular data processing. The Luminesce Alliance – Innovation for Children’s Health, is a not-for-profit cooperative joint venture between the Sydney Children’s Hospitals Network, the Children’s Medical Research Institute, and the Children’s Cancer Institute; It has been established with the support of the NSW Government to coordinate and integrate pediatric research; Luminesce Alliance is also affiliated with the University of Sydney and the University of New South Wales Sydney (2018–22). The Medical Research Future Fund, Australian Brain Cancer Mission /National Health & Medical Research Council/Lifting Clinical Trials and Registry Capacity (NHMRC MRF9500002), the Minderoo Foundation’s Collaborate Against Cancer Initiative (2018-2020) and funds raised through the Zero Childhood Cancer Capacity Campaign (2016-2020), a joint initiative of Children’s Cancer Institute and the Sydney Children’s Hospital Foundation, supported the national clinical trial and associated clinical and research personnel. G.M.M is funded by a Cancer Institute NSW Translational Program Grant, National Health and Medical Research Council (NHMRC) Synergy Grant and the Steven Walter Children’s Cancer Foundation. The KiCS program is supported by the Garron Family Cancer Centre at The Hospital for Sick Children through funding from the SickKids Foundation. A.S. is partially supported by a Garron Family Chair in Childhood Cancer Research; funding from The Canadian Institutes of Health Research; and National Cancer Institute of the National Institutes of Health, award number 5R03CA283675.

## Author contributions

M.H., G.M.M. and D.S.Z. conceived ZERO. V.T. is program director of ZERO. A.V. and A.S. co-direct KiCS. F.A., A.A., K.B., M.C., S.F., N.G.G., M.K., S.L.K., N.M., G.M., A.S.M., W.N., M.O., B.P., A.L.R., L.S. and P.J.W. were site principal investigators and site clinicians. J.D., C.D., A.J.G. and M.R. provided pathology oversight. M.W., C.M., S.E.-K., L.C., A.L., P.P., S.D., K.E.Y., M.P., A.S. and M.J.C. led the bioinformatic and computational genomics analyses. D.-A.K.-Q., A.V., L.M.S.L., P.B., A.-K.A., M.R., K.D. and C.M. analyzed and curated molecular findings. D.-A.K.-Q., L.M.S.L., T.N.T., G.M.M. and D.S.Z. led the MTB discussions and recommendations. A.J.S., D.-A.K.-Q., A.V., L.M.S.L., N.A.F.-B., E.K.C., N.S., S.J., S.S. and K.M.T conducted the clinical curation, MTB presentations and reports. A.J.S., D.-A.K.-Q., A.V., S.T., N.S., J.S., S.J. and S.S collected the clinical data. A.J.S., D.- A.K.-Q., A.V., M.J.C. and D.S.Z conceived, designed, analyzed and wrote the manuscript, with contributions and review from all authors.

## Extended Data Figures

**Extended Data Fig. 1:**
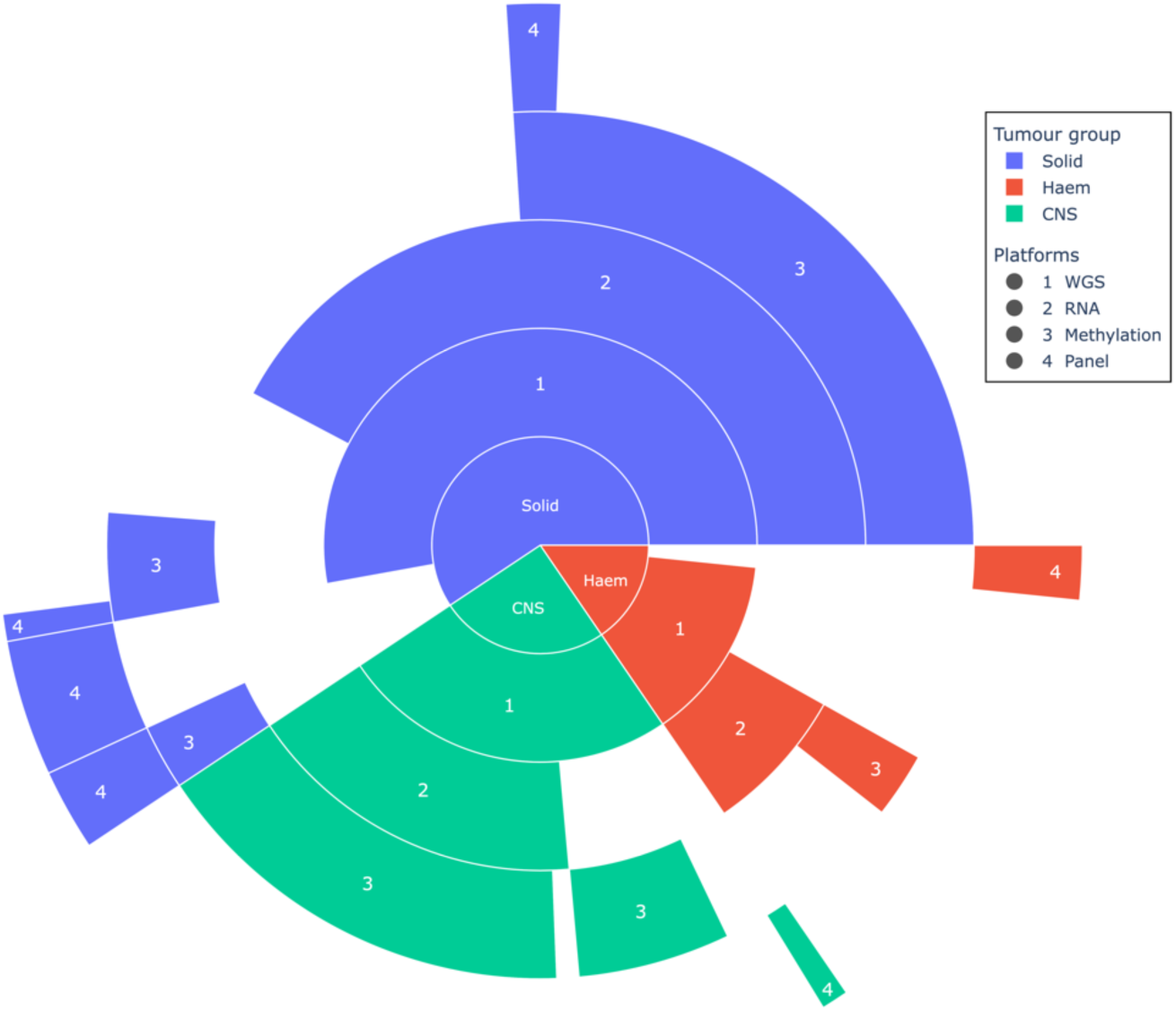
Sequencing Approaches in the Study Cohort. Sunburst plot showing genomic profiling strategies for the 123 patients. Sample submission influenced the type of genomic profiling performed with 66.7% (82/123) undergoing paired tumor-germline WGS and WTS (with two of these patients also receiving tumor panel sequencing), 25.2% (31/123) undergoing paired tumor-germline WGS alone (with two of these patients also receiving panel sequencing) and germline WGS performed in 8.1% (10/123) of patients for whom only FFPE tumor samples were available. In addition, DNA methylation profiling was performed in patients with CNS tumors, suspected sarcomas and selected other tumors with an adequate sample, comprising 56.9% of the cohort (70/123).

**Extended Data Fig. 2:**
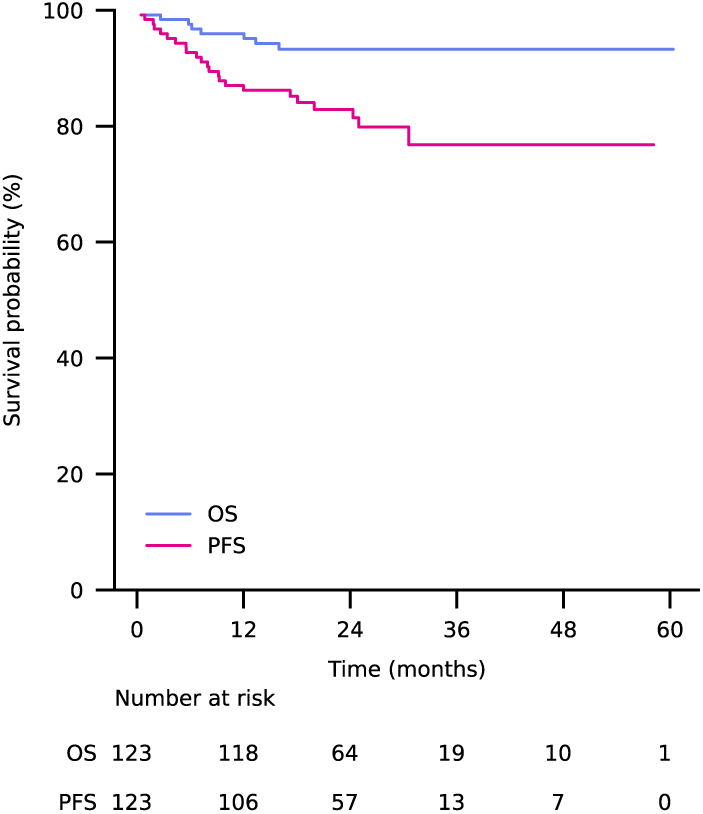
Kaplan–Meier Survival Analysis. Kaplan-Meier survival curve of the entire cohort. PFS was defined as the time from enrolment to disease progression, recurrence, or death from any cause, whichever occurred first. Abbreviations: OS, overall survival; PFS, progression free survival.

**Extended Data Fig. 3.**
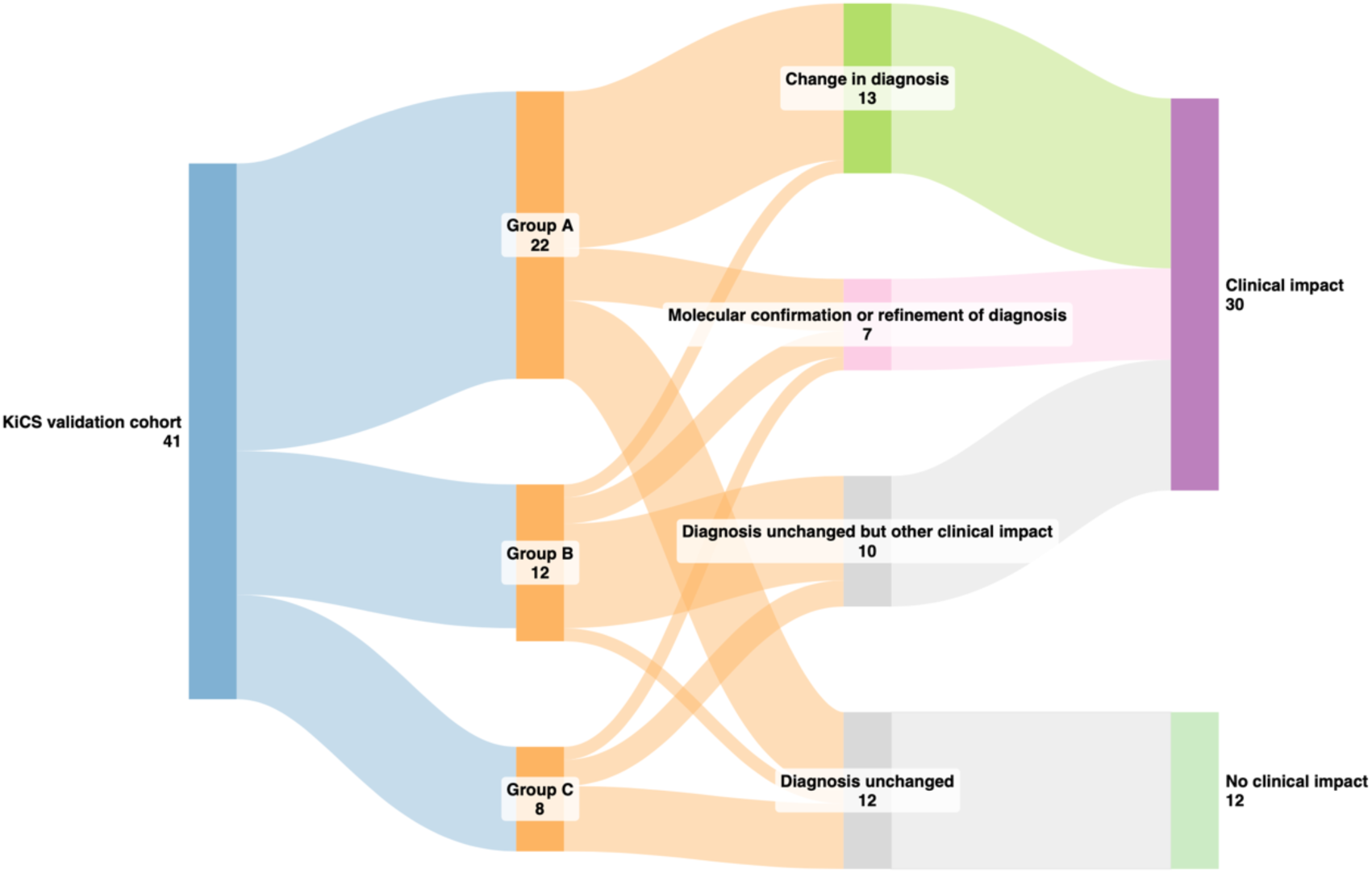
Validation of Comprehensive Genomic Profiling Outcomes in a Canadian Cohort. A Sankey diagram of the 41 patients eligible for outcome analysis from the KiCS cohort, treated here as an independent validation cohort from the KiCS precision medicine program led by The Hospital for Sick Children (SickKids), Canada. Clinical impact was defined as diagnostic utility, impact on conventional oncology management, precision guided therapy (PGT) recommendation and/or treatment, prognosis, identification of an underlying pathogenic or likely pathogenic germline variant not previously known, and/or other clinician-valued benefit.

## Extended Data Tables

**Extended Data Table 1: Baseline demographics.** Abbreviations: CNS, central nervous system; LGG, low grade glioma.

**Extended Data Table 2: Definitions of change in diagnosis, molecular clarification, molecular refinement, clinical impact categories.**

**Extended Data Table 3: Genomic findings and clinical impact of patients with formal change in diagnosis.**

**Extended Data Table 4: Validation cohort characteristics.**

## Supplementary Tables

**Supplementary Table 1: Reportable genomic findings**

